# Systematic Exploration of Hospital Cost Variability: A Conformal Prediction-Based Outlier Detection Method for Electronic Health Records

**DOI:** 10.1101/2025.01.10.25320349

**Authors:** François Grolleau, Ethan Goh, Stephen P. Ma, Jonathan Masterson, Ted Ross, Arnold Milstein, Jonathan H. Chen

## Abstract

Marked variability in inpatient hospitalization costs poses significant challenges to healthcare quality, resource allocation, and patient outcomes. Traditional methods like Diagnosis-Related Groups (DRGs) aid in cost management but lack practical solutions for enhancing hospital care value. We introduce a novel methodology for outlier detection in Electronic Health Records (EHRs) using Conformal Prediction. This approach identifies and prioritizes areas for optimizing high-value care processes. Unlike conventional predictive models that neglect uncertainty, our method employs Conformal Quantile Regression (CQR) to generate robust prediction intervals, offering a comprehensive view of cost variability. By integrating Conformal Prediction with machine learning models, healthcare professionals can more accurately pinpoint opportunities for quality and efficiency improvements. Our framework systematically evaluates unexplained hospital cost variations and generates interpretable hypotheses for refining clinical practices associated with atypical costs. This data-driven approach offers a systematic method to generate clinically sound hypotheses that may inform processes to enhance care quality and optimize resource utilization.

## Introduction

Marked variance amongst inpatient hospitalization costs impedes the efficient allocation of scarce medical resources and adversely affects healthcare quality, patient outcomes, and medical charges. In the United States, hospital costs remain significantly high, posing a substantial financial burden on both healthcare institutions and patients. The widespread use of Diagnosis-Related Groups (DRGs) has provided a foundational methodology for categorizing hospital cases to support cost management (1). However, accurately predicting and modulating hospital costs remains challenging. This challenge is further complicated by variations attributable to uncontrollable patient factors, such as illness severity or comorbidities present at admission, as well as deviations in medical practice and care delivery.

A significant amount of current research and operational efforts, by organizations such as Vizient, have focused on predictive modeling techniques. These techniques identify risk factors associated with adverse outcomes, like increased mortality or length of stay, and play a critical role in calculating observed-to-expected cost ratios, thereby informing reimbursement strategies (2–5). Despite these advancements, there is a dearth of literature addressing the modifiability of hospital costs (6). Much work has illuminated predictability, but the modifiability of medical costs due to care delivery practices remains underexplored.

Estimating causal effects using Electronic Health Records (EHRs) is particularly challenging due to the reliance on observational data and the dynamic nature of care delivery. Each potential medical intervention would typically require expert knowledge to specify the variables causing each intervention at different points in a patient’s trajectory (7), which is impractical given the vast number of interventions to evaluate. Without a systematic approach, numerous plausible areas for quality and efficiency improvements in healthcare—such as the early discharge process, nursing home placement accessibility, goals of care conversions, deterioration detection, nutrition, physical therapy early mobility, weekend discharge practice, bowel regimen, adequate hydration and electrolyte management, and transfusion practices—remain speculative.

In this paper, we introduce a novel methodology using Conformal Prediction-based outlier detection for EHRs to systematically generate and prioritize hypotheses about high-value care processes and areas for improvement in hospital practice. Similar to Phenome-Wide Association Studies (8) (PheWAS), which associate genetic variants with a wide range of phenotypic traits, our approach systematically identifies and prioritizes care processes. While most predictive modeling studies do not quantify uncertainty in their predictions, we leverage Conformal Quantile Regression (CQR), a state-of-the-art framework, to provide valid prediction intervals for any machine learning model (9). To the best of our knowledge, this study is the first to apply conformal inference as an outlier detection method for EHR data, despite its prior use in other domains (10).

Additionally, integrating Conformal Prediction in cost variability analysis facilitates the re-engagement of human experts in the loop (11). This method enables healthcare professionals to uncover insights into specific care processes that can be optimized, facilitating a more data-informed and expert-guided approach to enhancing hospital cost efficiency and patient care outcomes.

Our objective is to develop a method for systematically evaluating unexplained hospital cost variability and to generate plausible, interpretable hypotheses for clinical care practice patterns associated with higher or lower-than-expected costs. This approach aims to provide actionable insights that can improve care quality and efficiency in clinical operations.

## Methods

An overview of our workflow for systematic exploration of hospital cost variability is shown in Figure 1.

**Figure 1.**
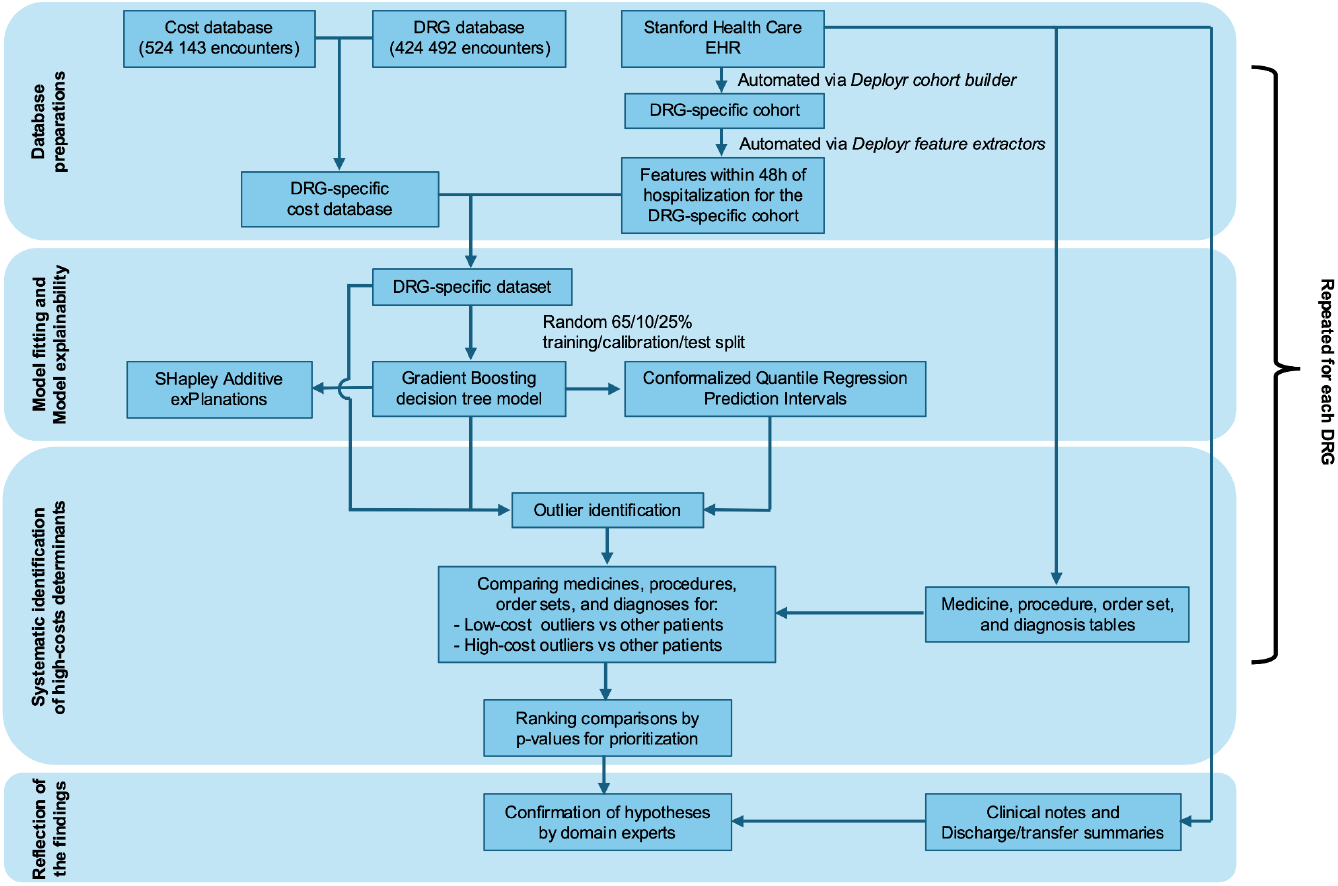
The conformal prediction-based outlier detection workflow for systematic exploration of hospital cost variability. Methodological details are provided in the body text.

### Data source and database preparations

The data consists of deidentified Electronic Health Records (EHR) for 33 077 inpatients treated at a large academic medical center, Stanford Health Care in Palo Alto, CA, between March 2019 and August 2021. These records are obtained by merging the cost and Diagnosis-Related Group (DRG) databases, incorporating both MS-DRG and APR-DRG classification systems. The merging process utilizes patient identifiers along with admission and discharge dates.

Unless otherwise mentioned all following steps are repeated separately for each DRG. For each patient-hospital encounter, we link the cost and DRG databases. Using the Deployr cohort builder (12) on Stanford Health Care EHR, we create DRG-specific cohorts. We subsequently apply the Deployr feature extractor pipeline (12) onto these cohorts to extract clinical features within 48 hours of hospitalization. Given that this automated feature extraction pipeline produces sparse, high-dimensional feature matrices, we concentrate on the two hundred features with the least amount of missing data. The resulting DRG-specific datasets include clinical features such as diagnosis (ICD-10) codes, medication orders, demographic variables, and laboratory results.

### Model fitting

In our approach, for each DRG-specific dataset, we build a model to predict the direct inpatient costs (scaled relative to the average inpatient admission cost) from the features available within 48 hours of hospitalization. For scalability without hurting models’ predictive abilities (13) missing data is managed through imputation by the mode. Indicator variables denoting missingness are concatenated to the imputed feature matrices. The datasets are randomly split into training, calibration (for conformalization, *see below*), and test sets with a 65/10/25% ratio. All models are gradient-boosting decision trees (14,15), which have proven highly effective for tasks involving tabular data (16,17). Model’s hyperparameters determined by minimizing the 5-fold cross-validated mean absolute error through randomized search (18). Models’ predictive performance is evaluated on held-out test sets by calibration plots including a smoothed (spline-based) calibration curve as well as the R^2^, Root Mean Squared Error (RMSE), intercept, and slope metrics (19).

### Model explainability

To gain a first line of insights into the determinant driving high (and low) hospital costs, we calculate SHappley Additive exPlanation values (SHAP) (20) from our models for all observations in a DRG-specific dataset. Feature importance is systematically assessed via Beeswarm plots.

To explore uncertainty in predicted costs, we calculate prediction intervals (PIs) for all predictions from our models using conformalized quantile regression (CQR) (9,21). Conformal prediction is a distribution-free uncertainty quantification method that has finite-sample guarantees applying to any non-parametric machine learning model, including gradient-boosting decision trees. In practice, we rely on the MAPIE library (22) and the held-out calibration sets to conformalize the standard (ie biased) PIs from quantile regression. To investigate cost variability, for each DRG, we calculate the mean for CQR PIs’ (i) lower bound, (ii) upper bound, and (iii) widths, all categorized by fifths of predicted costs.

### Outlier identification

We characterize as high- (low-) cost outliers the patients whose observed cost was above (below) the upper (lower) bound of their 50% CQR PI. The nominal coverage of the PIs can be adjusted to vary the outlier group size. In our experiments, 50% PIs provide a reasonable balance between outlier group size and mean difference in costs for outliers vs other patients.

### Systematic identification of high- (low-) cost determinants

For each DRG, we use Stanford Health Care EHR to compare the medicines, procedures, order sets, and diagnoses associated to high-cost outliers vs other patients. These comparisons are repeated for low-cost outliers vs other patients. More precisely, we query the medicine, procedure, order set, and diagnosis tables from Stanford Health Care EHR for outliers and nonoutlier patients, for all DRGs. The exposure of outliers to various medicines, procedures, or order sets, is compared to the corresponding exposure in nonoutliers. We build contingency tables for all comparisons, and the differences in exposures are quantified through odds ratios and p-values from Fisher exact tests.

To facilitate prioritization, we merge results across all DRGs, for each exposure comparison (medicines, procedures, order sets, and diagnoses) and remove duplicates. Lastly, mimicking the methodology from PheWAS studies (8), we rank all comparisons by ascending p-values to highlight the most promising hypotheses for cost reduction. Since p-values are used exclusively for ranking associations, we do not apply corrections for multiple comparisons. Therefore, the magnitude of the p-values should be interpreted as exploratory and hypothesis-generating.

### Reflection of the findings

In the final step, we engage domain experts—physicians, internal medicine specialists, hospitalists, and hospital finance specialists (specifically FG, EG, JHC, and JM in this study)—to evaluate the prioritized cost association hypotheses. The subject experts then review these to assess which medicines, procedures, and order sets could imply plausible interventions/scenarios/modifiers for improving value without compromising health outcomes.

## Results

### Model evaluation and explainability

Our workflow produces calibration plots, evaluation metrics, SHAP values, and cost variability assessment for each DRG-specific prediction model. As an example, in Figure 2, we show the output of our pipeline for the prediction model corresponding to the DRG “open craniotomy except trauma.”

**Figure 2.**
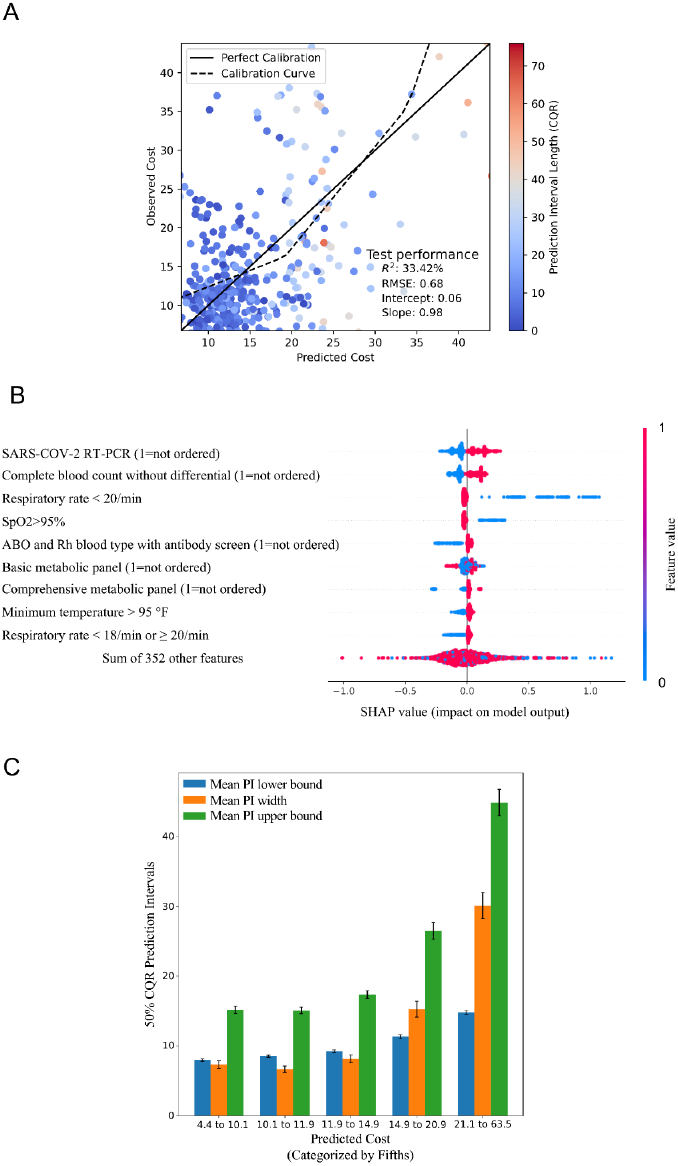
Example output from our systematic exploration pipeline for model fitting and explainability and the DRG “open craniotomy except trauma.”

Panel A: Evaluation of the DRG-specific gradient-boosted tree model on the held-out test set. On Panel A, each dot represents a patient and the dot’s color depicts the uncertainty (50% CQR PI width) in the prediction for that patient. A positive value of R^2^ indicates that the model using features collected within 48 hours of hospitalization, outperforms a model that would predict the mean cost for the relevant DRG.

Panel B: Feature importance for SHAP values. For a better understanding of our machine learning models, our workflow systematically showcases SHAP values in a Beeswarm plot. Panel B illustrates that the absence of COVID-19 PCR testing correlates with higher hospital costs. Additionally, missing other medical orders, including complete blood count, ABO/Rh testing, and metabolic panels, are also correlated with increased costs. Severity indicators such as respiratory rate, SpO2, and temperature exhibit complex associations with costs, which might not be easily discernible when considering these variables individually.

Panel C: Investigation of DRG-specific cost variability by fifths of the predicted costs. To investigate whether cost variability was greater in high or low-cost patients, we systematically represent uncertainty (50% CQR PI width) in the prediction categorized by fifths of predicted costs. The vertical intervals are ± standard errors for the means. Panel C shows that the uncertainty in the predicted cost was greatest in the patients with the highest predicted cost.

In all panels, costs are scaled relative to the average inpatient admission cost.

### Systematic identification of high- (low-) cost determinants

Our framework prioritizes hypotheses to improve the value of care by enabling comparisons of procedures, medications, order sets, and diagnoses associated with high- and low-cost outliers relative to other patients across all DRGs. For instance, in Table 1 we present comparisons for medical order sets where all top ten comparisons with the lowest p-values are related to high-cost outliers. Comparisons for low-cost outliers can be readily explored by conditioning on these comparisons. Table 2 provides an example of comparisons involving low-cost outliers. In both cases, we offer interpretations based on the review of clinical notes and transfer summaries of a selection of outlier patients.

**Table 1.**
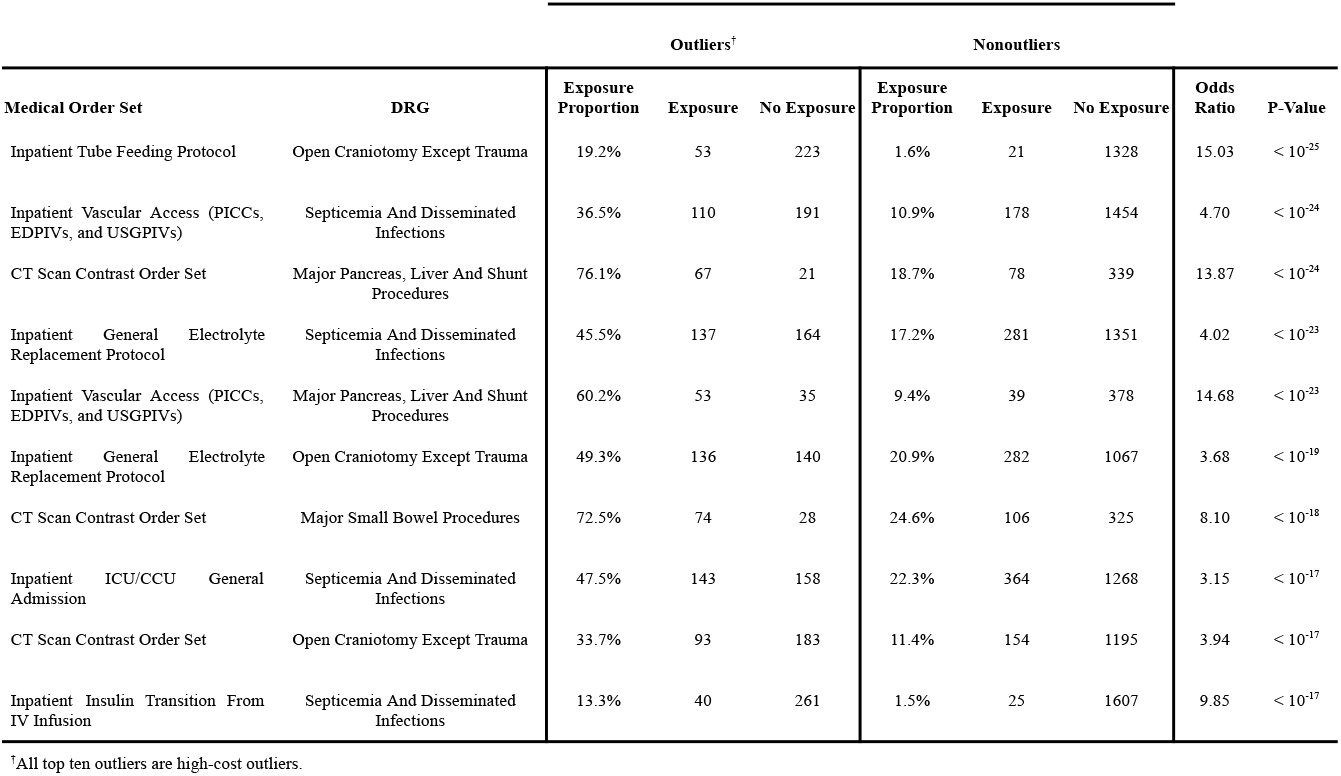
Top ten medical order sets entries with outliers vs nonoutliers comparisons ranked by p-values for prioritization. The outliers versus nonoutliers patients are identified by using conformal inference. The “Exposure” and “No Exposure” columns denote the number of patients within a specific DRG who received or did not receive the corresponding medical order set, respectively. The proportions in each category are calculated as 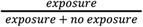. The odds ratios are computed as 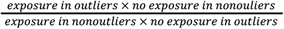. An odds ratio greater than 1 indicates a positive association between the medical order set and the presence of high-cost outliers. Patients undergoing “Open Craniotomy Except Trauma” who receive the “Inpatient Tube Feeding Protocol” incur higher than expected costs, likely because those patients needing specialized nutritional support during recovery require more extensive and lengthy care. Similarly, those with “Septicemia and Disseminated Infections” needing “Inpatient Vascular Access (PICCs, EDPIVs, and USGPIVs)” likely require extended intravenous therapy, indicating severe illness and higher costs to administer treatment. Procedures like “Major Pancreas, Liver, and Shunt Procedures” and “Major Small Bowel Procedures” using the “Radiology CT Contrast Order Set” necessitate detailed imaging, indicating greater complexity and expense in their management. The “Inpatient General Electrolyte Replacement Protocol” for conditions like “Septicemia and Disseminated Infections” and “Open Craniotomy Except Trauma” suggests correction of critical imbalances, reflecting higher illness severity. The “Inpatient ICU/CCU General Admission” and “Inpatient Insulin Transition From IV Infusion” for critically ill patients point to extensive care and resource use, leading to higher costs. These findings provide assurance that we are not identifying false correlations while gaining deeper insights into how tube feeding, vascular access needs, and insulin drip requirements stand out as being more strongly associated with higher-than-expected costs. CT=Computed Tomography. DRG=Diagnosis-Related Group. EDPIV=Emergency Department Peripheral Intravenous Line. PICC=Peripherally Inserted Central Catheter. USGPIV=Ultrasound-Guided Peripheral Intravenous Line.

**Table 2.**
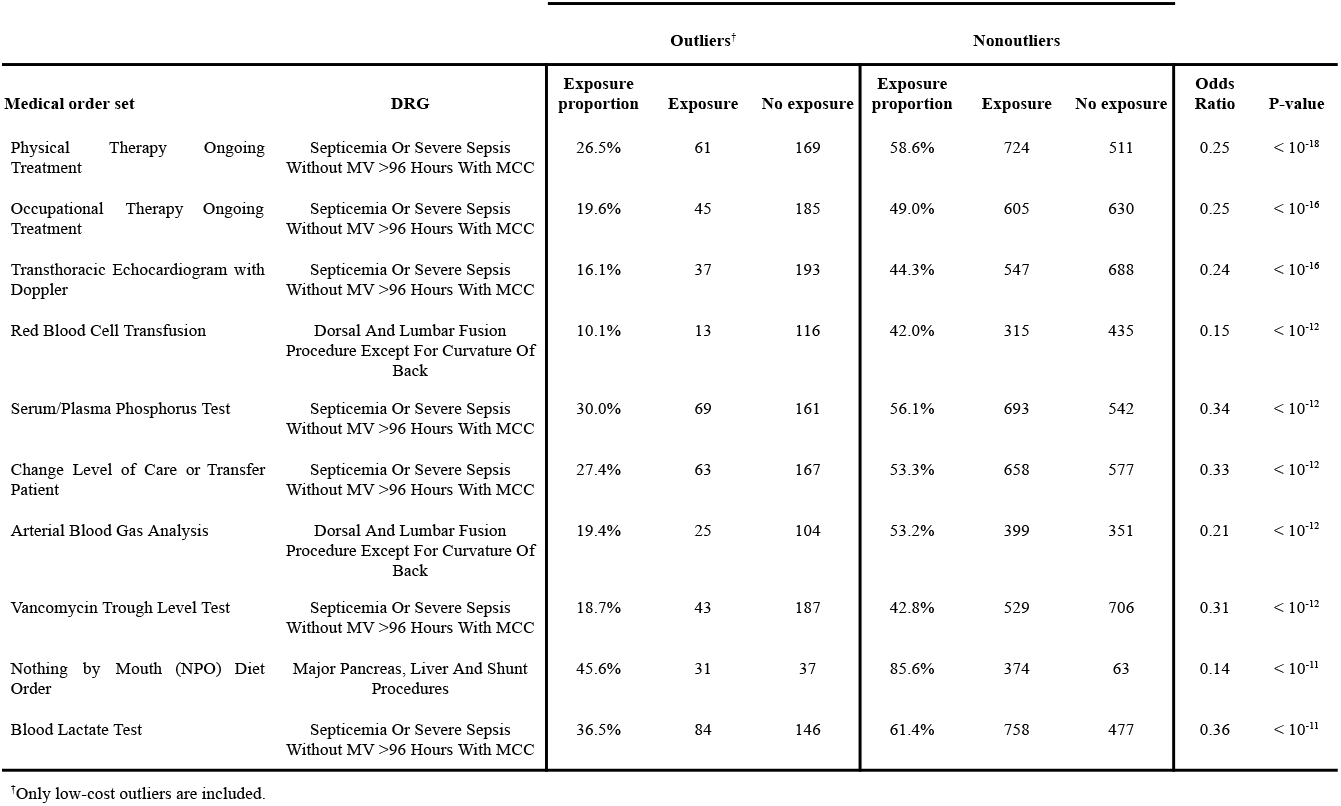
Top ten medical order sets entries for low-cost outliers. The comparisons between low-cost outliers vs nonoutliers are ranked by p-values. The outliers versus nonoutliers patients are identified by using conformal inference. The “Exposure” and “No Exposure” columns denote the number of patients within a specific DRG who received or did not receive the corresponding medical order set, respectively. The proportions in each category are calculated as 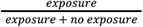. The odds ratios are computed as 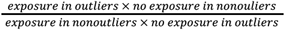. An odds ratio lower than 1 indicates a positive association between the medical order set not being received and the presence of low-cost outliers. Patients with “Septicemia Or Severe Sepsis Without MV >96 Hours With MCC” who do not receive treatments such as “Physical Therapy,” “Occupational Therapy,” and various diagnostic tests have lower-than-expected hospital costs, suggesting less severe conditions. Similarly, those undergoing “Dorsal and Lumbar Fusion” without the need for “Red Blood Cell Transfusion” or “Arterial Blood Gas Analysis,” as well as “Major Pancreas, Liver, and Shunt Procedures” patients not on an “NPO Diet Order,” also incur lower-than-expected costs, indicating simpler clinical situations. These findings confirm that we are not detecting spurious correlations and provide deeper insights into how physical therapy and parenteral nutrition, rather than other order sets, are more negatively associated with lower-than-expected costs. DRG=Diagnosis-Related Group. MV=Mechanical Ventilation. MCC=Major Complications or Comorbidities. NPO=*Nil Per Os*.

## Discussion

In this analysis of Stanford EHR data, we have applied a systematic method to generate clinically sound hypotheses aimed at enhancing care quality and optimizing resource utilization. The originality of our approach lies in the integration of conformal inference with traditional machine learning techniques to identify high-cost and low-cost outliers. Our analysis revealed that interventions such as tube feeding, vascular access needs, insulin drip, physical therapy, and parenteral nutrition stand out as being more strongly associated with higher-than-expected costs. If validated, these findings suggest that focusing quality and efficiency improvements on these interventions may enhance the value of inpatient care.

Beyond the specific results observed at Stanford, our approach holds promise for broader applications. The systematic integration of conformal inference with machine learning can be implemented in other healthcare institutions to uncover and prioritize areas where care processes have higher-than-expected costs. By identifying cost outliers, these institutions can pinpoint specific procedures or diagnoses that require targeted interventions to improve efficiency. This flexible and adaptable approach provides a robust framework for continuous improvement across a variety of clinical settings, ensuring that health care systems can optimize costs while maintaining or enhancing care quality.

However, our analysis had several limitations. Within certain DRGs, such as “heart failure” and “respiratory infections,” cost prediction models demonstrated limited performance on left-out test sets. This limitation is mitigated by the primary objective of our method, which is to retrospectively identify high-cost and low-cost outlier patients rather than develop models for prospective deployment.

Additionally, the data was collected between March 2019 and August 2021, during the COVID-19 pandemic. Rapidly evolving clinical practices during this period may have impacted our hypothesis generation and prioritization. Despite this, we successfully generated clinically sound hypotheses, validated by experts. We anticipate our methodology to be more robust when clinical practices are stable over time.

As in PheWAS studies, our method ranks associations rather than evaluating causal effects. This scalability requires human expertise at the final stage to confirm or refute the causal likelihood of the generated hypotheses. Enhancing this step with language models to extract relevant information from clinical notes and discharge summaries could improve efficiency. We are currently working on solutions to facilitate this process.

The American public and policymakers have made significant investments in digitizing health records. We have demonstrated proof of concept for our systematic method to generate clinically sound hypotheses that may inform processes to enhance care quality and optimize resource utilization. Other institutions can adopt our workflow to quantify cost inefficiencies associated with specific diagnoses or procedures and prioritize cost-reduction efforts by their potential impact.

## Conclusion

We present a proof of concept for integrating conformal prediction with machine learning to systematically explore hospital cost variability. By incorporating human experts back into the decision-making loop, this approach not only identifies areas for potential cost savings but also aims to maintain or enhance the quality of care. This methodology offers a valuable tool for healthcare systems seeking to optimize resource utilization and improve patient outcomes. Our findings highlight the potential for data-driven strategies to inform clinical practice and drive efficiency improvements in hospital settings.

## Data Availability

All data produced in the present study are available upon reasonable request to the authors.

## Contributions

FG: study ideation and design, analysis, interpretation of results, manuscript first draft; EG: study ideation and design, clinical interpretation; SPM: study ideation and design, clinical interpretation; JM: study ideation and design, operational interpretation; TR: provided the cost data, operational interpretation; AM: study ideation, curated and provided data; JHC: study ideation and design, interpretation of results, manuscript first draft. All authors reviewed and provided feedback on manuscript.

## Acknowledgments

We thank Selina Pi and Conor K. Corbin for their helpful feedback on the hospital cost database and the Deployr feature extraction pipeline respectively. The authors report no disclosures or conflicts of interest.

